# Estimating the Global Target Market for Passive Chlorination

**DOI:** 10.1101/2022.10.27.22281472

**Authors:** Katya Cherukumilli, Robert Bain, Yiru Chen, Amy J. Pickering

## Abstract

Deployment of passive (in-line) chlorinators, devices that disinfect water without electricity or daily user input, is one strategy to advance access to safely managed drinking water. Using the Joint Monitoring Programme (JMP) data, we first calculate the number of people in low- and middle-income countries (LMICs) using drinking water sources that are either compatible (piped water, kiosks) or potentially compatible (packaged/delivered water, rainwater, tubewells, boreholes, protected springs) with passive chlorinators. Leveraging water quality data from the Multiple Indicator Cluster Surveys (MICS), we estimate that 2.32 [95% CI: 2.19, 2.46] billion people in LMICs use microbially contaminated (with fecal indicator bacteria) drinking water sources that are compatible (1.51 [1.42, 1.60] billion) or potentially compatible (817 [776, 858] million) with passive chlorinators. The largest target market for passive chlorinators is in South Asia (551 [532, 571] million rural users and 401 [384, 417] million urban users), where over 77% of drinking water sources compatible with passive chlorinators are contaminated. However, self-reported household water treatment practices indicate that chlorination is more common in the African and Latin American regions, suggesting passive chlorination would have higher acceptance in these regions compared to Asia. Reaching the full target market will require establishing passive chlorinator compatibility with handpumps and protected springs and identifying financially viable implementation models.

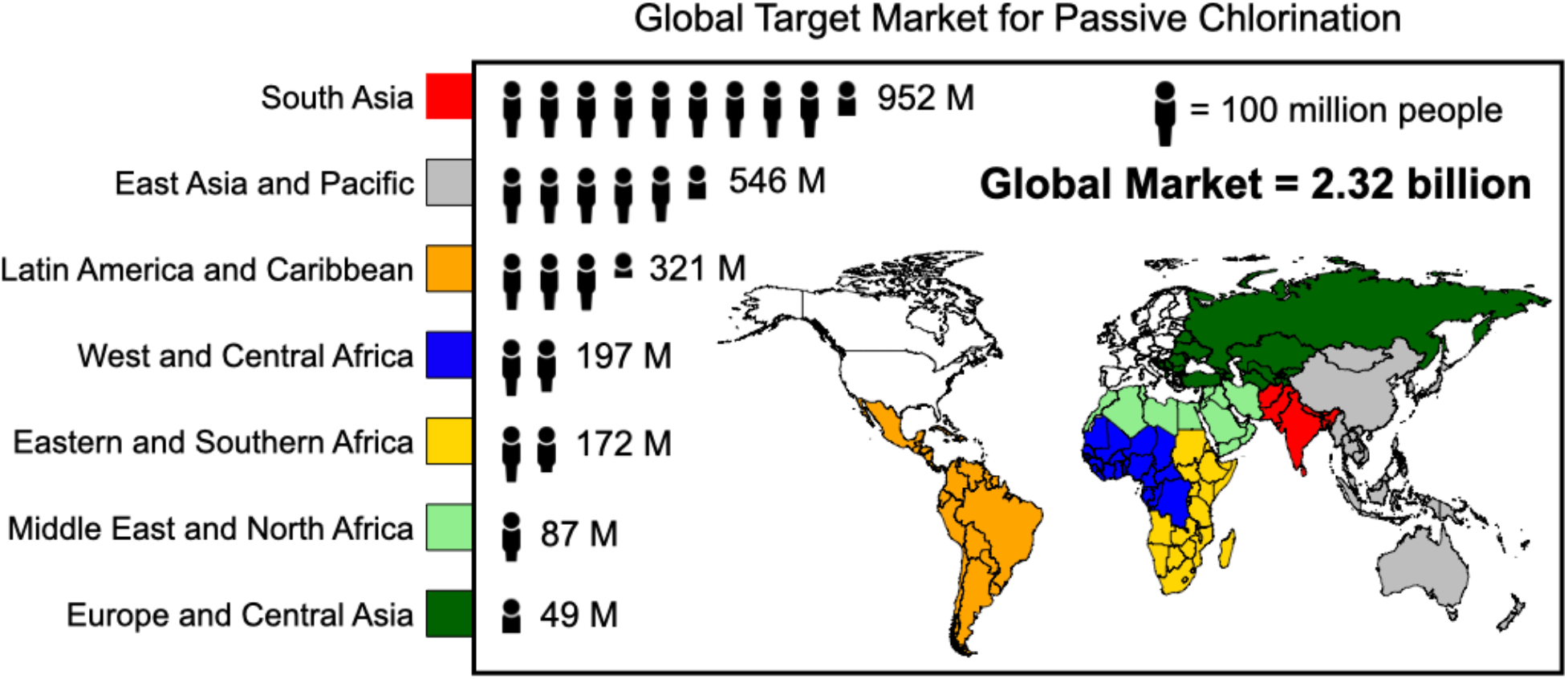

## 1. Introduction

The Joint Monitoring Programme (JMP) led by the World Health Organization (WHO) and United Nations Children’s Fund (UNICEF) tracks progress toward the UN Sustainable Development Goals targets 6.1 and 6.2, to provide drinking water, sanitation, and hygiene for all by 2030. Data collected by UNICEF’s Multiple Indicator Cluster Surveys (MICS) evaluate whether or not drinking water services used by households are “safely managed”, meaning the water is from an improved source, accessible on premises, available when needed, and free from contamination (see **Table S1** for additional details)^1^. Estimates for safely managed drinking water services have been reported for over 138 countries using over 3283 datasets for drinking water, including administrative data, population censuses, and household surveys including MICS^2^. In 2020, the JMP estimated that over 2 billion (>26%) people lacked access to safely managed drinking water services globally^2^.

Passive (in-line) chlorinator devices can potentially accelerate progress toward universal access to safely managed drinking water services^3,4^. Passive chlorinators offer the unique advantage of automatically and accurately dosing chlorine in water supplies in-line or at the point of collection, without requiring electricity, moving parts, or daily user input. Currently, the majority of commercially available passive chlorinators are compatible with piped water supplies or water storage tanks^5–14^. Ongoing research is evaluating the compatibility of passive chlorinators with manual handpumps^3,15,16^. Passive chlorinators are not compatible with water sources that are typically more turbid, such as surface water, without additional pretreatment steps such as coagulation and/or filtration. Implementation settings for chlorine dosers in low- and middle-income countries (LMICs) include regions with intermittent piped water supply, where drinking water is typically microbiologically contaminated at the point of collection due to insufficient chlorine dosing, leaky pipes, and negative pressure^17^.

The main objectives of our study are: (1) to characterize the proportion of household drinking water sources that are compatible or potentially compatible with passive chlorinators using the JMP database; and (2) to estimate global and regional target markets for passive chlorinators in LMICs using source microbial water quality data from the MICS database. In addition, we examine geographic patterns in household water treatment methods to better understand user acceptance of chlorine.

## 2. Methods

### 2.1. Data Sources, Categorization, and Analysis

JMP estimates and MICS data were used to characterize household drinking water facility types, microbiological source water quality, and self-reported household water treatment methods. Country-level JMP facility type estimates for populations using piped and non-piped improved sources in 135 LMICs were used to identify populations using drinking water sources compatible or potentially compatible with passive chlorinators across seven UNICEF programme regions **(Table S2)**. Drinking water sources were defined as “compatible” (piped water supplies, public taps, standpipes, and water kiosks), “potentially compatible” (tubewells, boreholes, rainwater, tanker-trucks, cart with small tanks, bottled water, sachet water, and protected springs), or “incompatible” (protected dugwells, unprotected dugwells, unprotected springs, surface water, and other or unclassified sources) with passive chlorinators^4^ (**Table S3**). Additional details on classification and analysis of data on drinking water facility types from the 2020 JMP WASH Database are described in the Supporting Information (**Table S3)**.

To assess the microbial quality of drinking water sources, we used data of *E. coli* measurements taken at the point of collection from a subset of households chosen to complete the water quality testing module in the MICS questionnaires^18^. These surveys were conducted across 37 countries from 2014 - 2021 (**Table S2)**. To identify patterns in household water treatment methods, we analyzed the responses from all surveyed households (self-reporting treatment) across 54 countries in MICS5 and MICS6 surveys conducted from 2013 - 2021 (**Table S2)**. Self-reported household drinking water treatment methods in the MICS database were categorized as boiling, adding bleach/chlorine, using a filter (ceramic, sand, or composite), settling, solar disinfection, cloth straining, or other methods. We were motivated to conduct this analysis to update prior estimates of household water treatment methods made by Rosa and Clasen in 2010.^19^

### 2.2 Estimation of Regional and Global Target Markets in LMICs

The JMP estimates the global population using piped, non-piped improved, and unimproved sources by weighting the average proportion of the households reporting the use of these facility types in urban and rural regions with their respective population numbers^20^. In this study, we used a similar methodology to estimate the regional and global target markets for passive chlorinators in LMICs. We defined the upper bound target market as the total population using microbially contaminated drinking water sources that are compatible or potentially compatible with passive chlorination, and the lower bound as those using contaminated compatible water sources. First, we used the JMP WASH country database to characterize the usage (and passive chlorinator compatibility) of different drinking water source types in urban and rural areas of UNICEF programme regions **(Table S3)**. Next, we used the MICS5 and MICS6 data to calculate the proportion of water sources contaminated with *E. coli* by source type in the UNICEF programme regions. Using these two estimates, we then calculated the proportion of microbially contaminated household drinking water sources that are compatible or potentially compatible with passive chlorinators. Finally, we calculate the regional and global target markets by multiplying our household proportion estimates by urban and rural country population data in each region^21^. 95% confidence intervals were calculated around the proportion point estimates for household water quality (i.e., proportion of each source type contaminated with *E. coli)* assuming an underlying binomial distribution and the uncertainty was propagated through the estimation of the target market.

## 3. Results and Discussion

### 3.1. Microbial Contamination of Household Drinking Water Sources by Facility Type

Households surveyed by MICS most commonly report using piped water, packaged and delivered water, or boreholes as their primary source of drinking water **(Fig. S1, Table S4)**. Kiosk usage is uncommon, and the reliance on protected springs, rainwater, and surface water is more common in rural areas than urban areas **(Fig. S1)**. The microbial (*E. coli*) contamination of household drinking water sources measured at the point of collection also varies based on source facility type, region, and urban vs. rural residence (**Fig. 1, Table S5)**. Piped water supplies are typically more contaminated in rural areas than urban areas across regions, with the exception of South Asia, where > 75% of piped supplies in both urban and rural areas are microbially contaminated, likely due to intermittent water systems (**Table S5**)^17^. The microbial contamination of packaged/delivered water varies greatly, from approximately 3.4% [95% CI: 0.1, 17.8] in rural Europe and Central Asia to 90.9% [58.7, 99.8] in rural Eastern and Southern Africa **(Table S5)**. Across regions, households reliant primarily on rainwater are exposed to high levels of microbial contamination (often > 80%) in both urban and rural areas.

### 3.2. Household Drinking Water Source Compatibility with Passive Chlorinators in LMICs

A majority of households across all UNICEF programme regions report using drinking water source types that are compatible or potentially compatible with passive chlorinators (**Fig. 2A, Table S6**). In large part, this widespread compatibility is a result of increases in piped water coverage globally in the last two decades since 2000, from 57% (3.5 billion people) to 65% (5.1 billion people)^2^.

**Figure 1.**
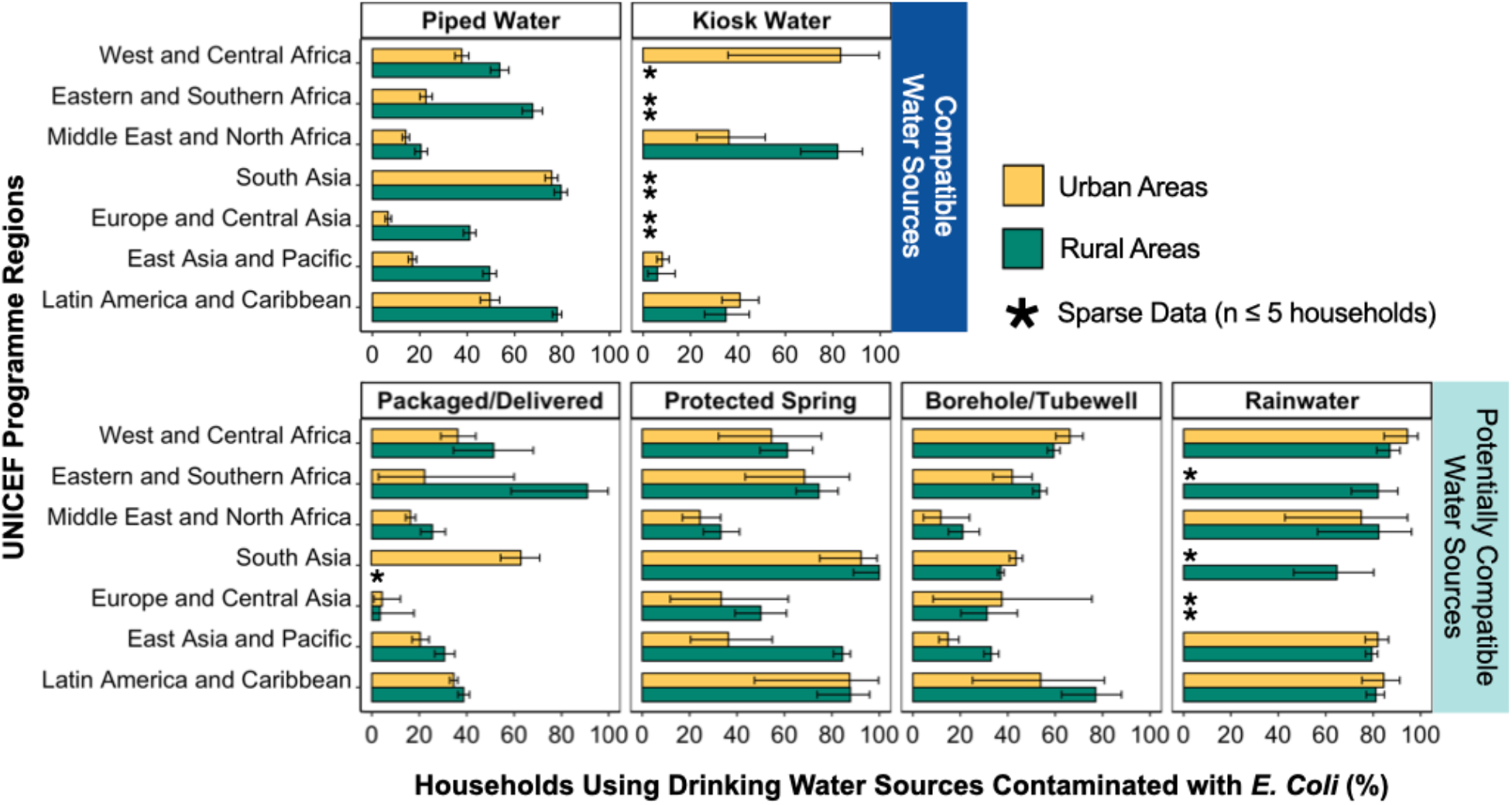
Microbial contamination of primary household drinking water sources, by facility type. Piped water and kiosk water are compatible with passive chlorinators and the remaining drinking water sources shown are potentially compatible (water sources incompatible with passive chlorination are not shown). Data on regional microbial (*E. coli*) water contamination in urban and rural households was obtained from water quality questionnaires in Multiple Indicator Cluster Surveys (MICS 5 & 6). Asterisks indicate regional locations where fewer than 5 households reported using a specific drinking water source facility. Error bars represent 95% confidence intervals calculated assuming an underlying binomial distribution. Full data is available in **Tables S5** in the Supporting Information.

**Figure 2.**
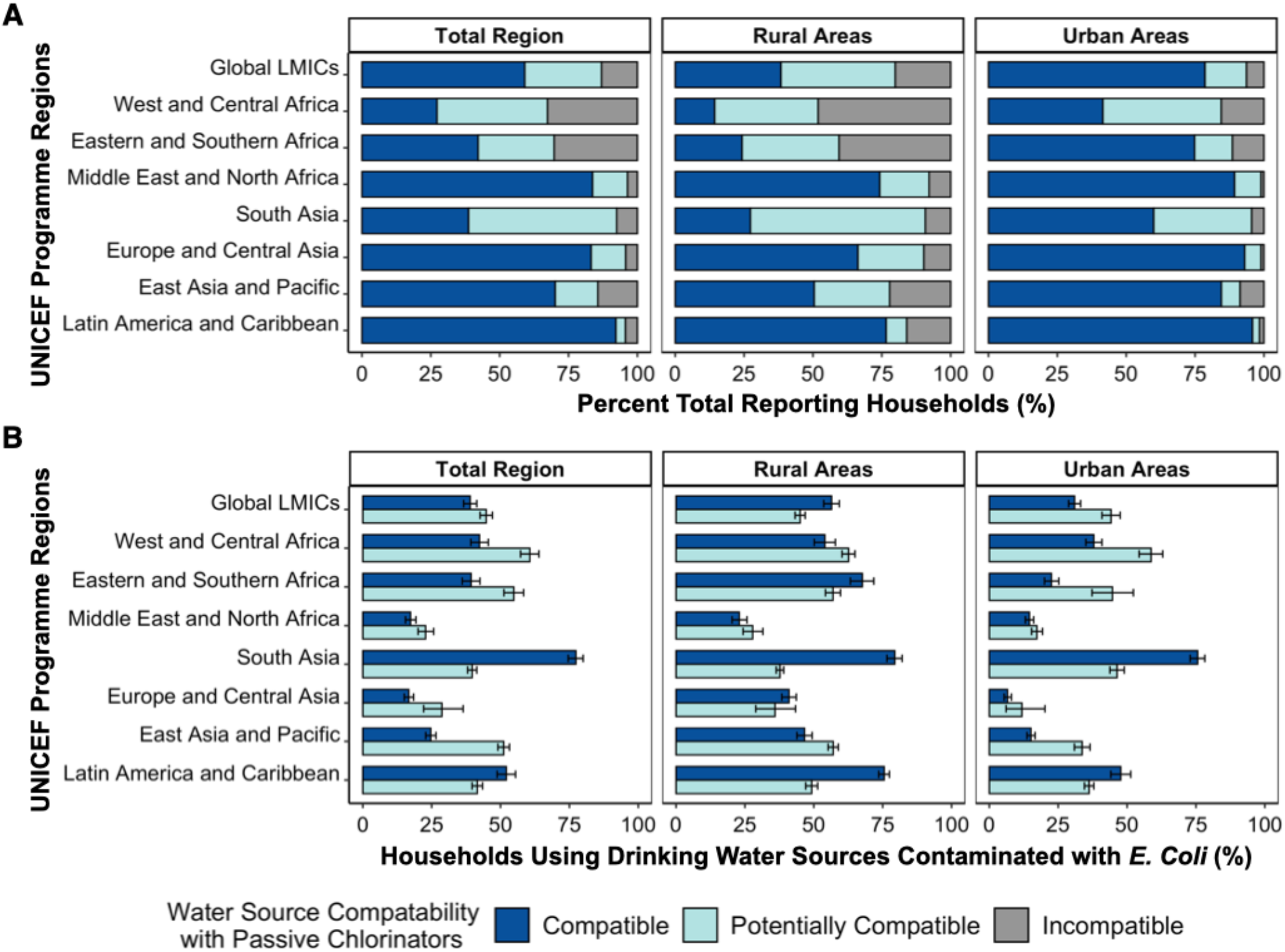
Household primary drinking water source compatibility with passive chlorinators (A), and (B) Percent (%) of compatible and potentially compatible water sources contaminated with *E. coli*. Water source categories are classified as being compatible in dark blue (piped supplies and water kiosks), potentially compatible in light blue (improved non-piped sources excluding protected dugwells), or incompatible in gray (unimproved sources and protected dugwells). Panel B data on regional microbial (*E. coli*) water contamination in urban and rural households was obtained from water quality questionnaires in Multiple Indicator Cluster Surveys (MICS 5 & 6). Error bars in panel B represent 95% confidence intervals calculated assuming an underlying binomial distribution. Additional data related to this figure can be found in **Tables S6, S7**, and **S8** in the Supporting Information.

In regions with lower piped water access (27.2 - 42.2% coverage in West and Central Africa; Eastern and Southern Africa; South Asia), a much larger proportion of urban households (41.5 - 74.8%) report using drinking water sources compatible with passive chlorinators in comparison to rural households (14.3 - 27.3%) (**Fig. 2A, Table S6)**. The largest disparity between urban and rural areas is evident in West and Central Africa and Eastern and Southern Africa, where over three-quarters of urban households (compared to less than roughly half of rural households) in the regions report using either compatible or potentially compatible sources. Particularly in rural South Asia, where borehole usage is most dominant worldwide, most households (63.5%) use potentially compatible water sources (**Fig. S1, Fig. 2A, Table S6)**.

Microbial contamination prevalence in household drinking water sources classified by compatibility is shown in **Figure 2B**. South Asia has the highest regional proportion (77.4% [95% CI: 74.6, 80]) of drinking water sources that are microbially contaminated and compatible with passive chlorinators (**Fig. 2B, Table S7)**. Across the other UNICEF programme regions, except for South Asia, higher levels of microbial contamination are consistently observed in compatible drinking water sources used in rural households (22.9 - 75.5%) compared to urban households (6.6 - 47.7%).

### 3.3. Regional Target Markets for Passive Chlorination in LMICs

The estimated total global target market for passive chlorinators in LMICs ranges from 1.51 [95% CI: 1.42, 1.60] billion compatible water source users to 2.32 [2.19, 2.46] billion compatible *and* potentially compatible source users (**Fig. 3, Table S9**). Within the market of compatible users, more people live in urban areas (819 [762, 877] million) than in rural areas (688 [653, 722] million). For the upper bound market, which includes potentially compatible water source users, the reverse is true: more people live in rural areas (1.28 [1.22, 1.34] billion) than in urban areas (1.04 [0.97, 1.12] billion).

**Figure 3.**
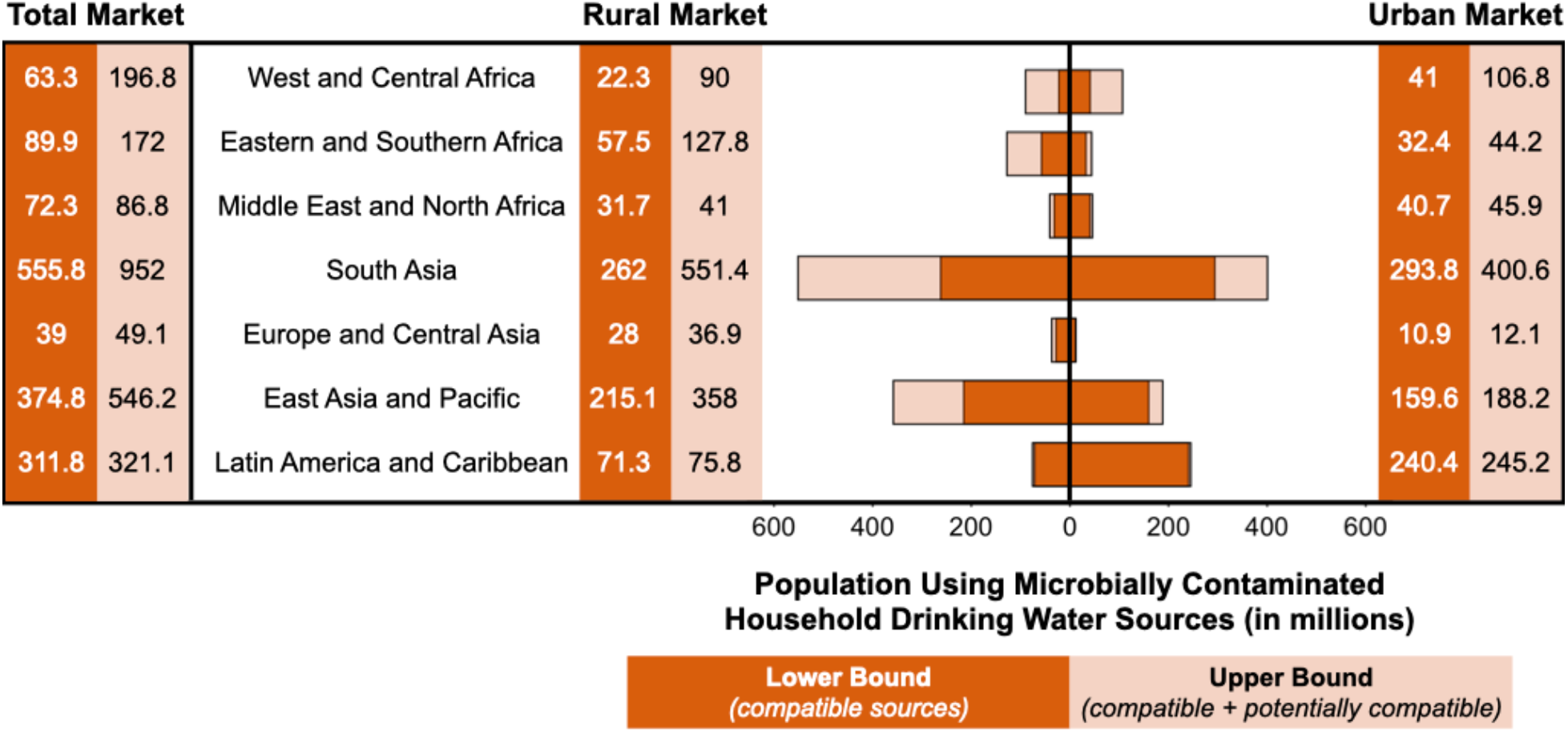
Regional target markets for passive chlorination, reported in millions of people using microbially contaminated water sources that are compatible (lower bound market) or compatible and potentially compatible (summed to form the upper bound market estimate). The total rural and urban population estimates for each region were calculated using the JMP 2021 and UNDESA population databases. Full data is available in **Table S9** in the Supporting Information.

The largest populations using contaminated drinking water sources that are compatible or potentially compatible with passive chlorination are in South Asia (952 [915, 988] million, majority rural users) and East Asia and Pacific (546 [511, 582] million, majority rural users). The overall target market range is largest in South Asia due to the high rates of microbial contamination of drinking water sources in the region (**Fig. 1** and **Fig. 2B**). Despite the highest rates of piped water coverage being in Europe and Central Asia (**Fig. 2A, Fig. S1)**, this region has the smallest target market bounds for passive chlorinators (39.0 [35.1, 43.1] million to 49.1 [42.9, 55.9] million users) due to its lower population in comparison to other UNICEF programme regions.

The global target market estimate presented here is conservative (i.e., an underestimate) due to the following reasons: (1) we only included LMICs, but there is also likely a target market for this technology in underserved settings in high-income countries; (2) water sources that tested clean at one time point could still be contaminated at other times and therefore would benefit from passive chlorination; and (3) water sources that tested clean at the point of collection often deteriorate in quality at the point of use^1^ and residual chlorine can offer the benefit of additional protection. In addition, our results rely on the assumption that the contamination levels for each water source type are representative of all countries in that region. The regional proportion and population estimates made in this current analysis are constrained by the lack of representative water quality data for populous countries including China, India, and Indonesia. Additionally, our analysis of the global and regional target markets for passive chlorination does not consider the ease of chlorine procurement, strength of local material supply chains, or device manufacturing capabilities^4^. In addition, we did not consider local and regional variations in the occurrence of aqueous chemical contaminants (e.g., arsenic, fluoride, etc.)^22^, which are not removed by passive chlorination. Additional limitations of our analysis and potential sources of bias are further described in **Table S10** in the Supporting Information.

### 3.3. Self-reported Household Drinking Water Treatment Practices

The most common household water treatment methods used in LMICs are boiling, filtration, and/or chlorination and the least common method is solar disinfection (**Fig. 4, Table S11)**. The proportion of households that reported boiling their water ranged widely, from < 2% in West and Central Africa and Middle East and North Africa to 36.4% [36.0, 36.8] in rural East Asia and Pacific. This trend is consistent with a study conducted by Rosa and Clasen in 2010, which analyzed 67 national surveys and found that boiling was the most dominant household water treatment method used by approximately 21% of study households^19^.

**Figure 4.**
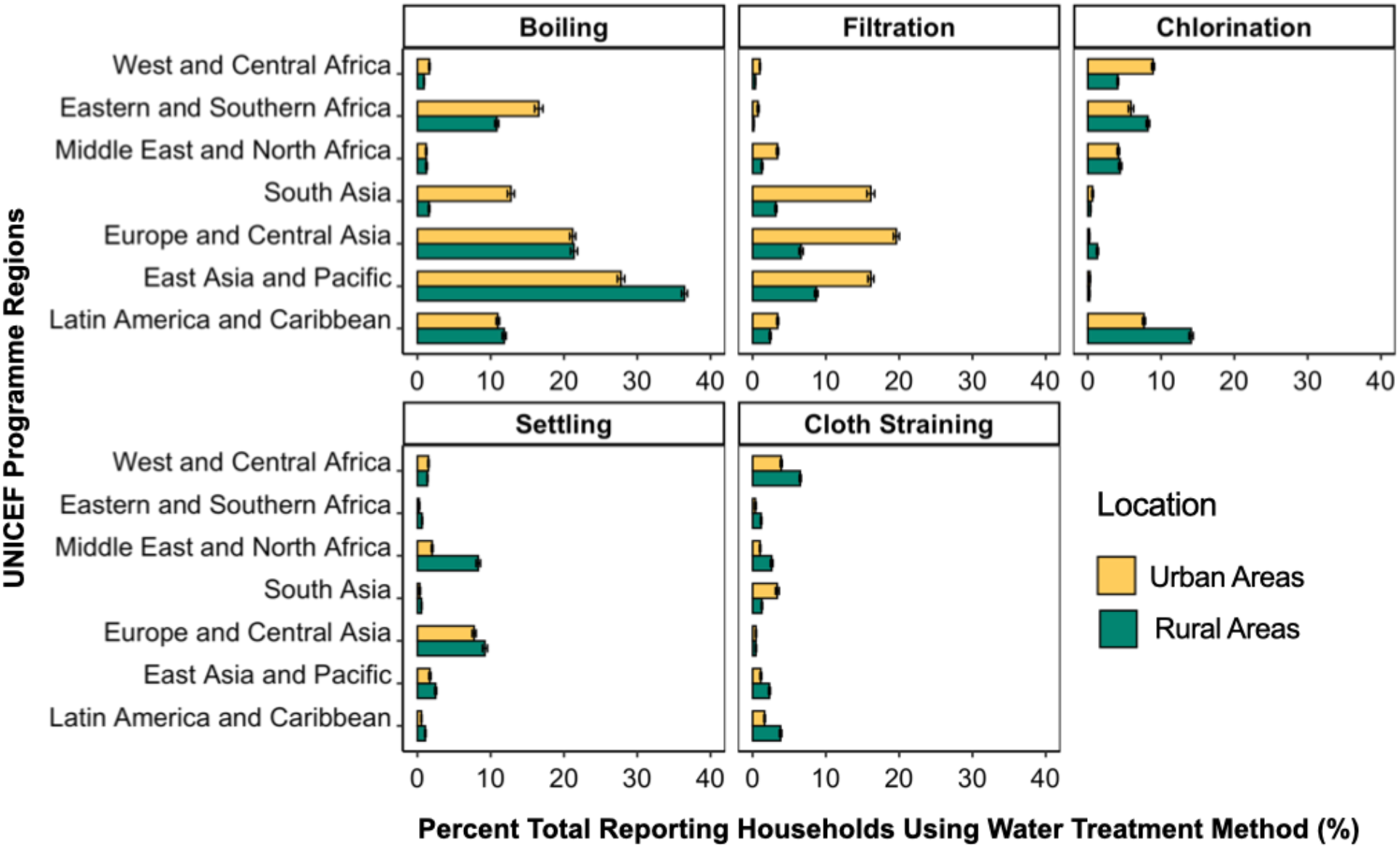
Household drinking water treatment practices, reported as percent (%) of total urban and rural households. Households could report multiple methods used including boiling, filtration (ceramic, sand, or composite filters), chlorination, settling, and/or cloth straining. Responses with proportions below < 1 % across regions (e.g., solar disinfection) are not shown. Error bars represent 95% confidence intervals calculated assuming an underlying binomial distribution. The total number of households in each region surveyed are as follows: West and Central Africa (216,419), Eastern and Southern Africa (68,056), Middle East and North Africa (87,161), South Asia (73,862), Europe and Central Asia (65,646), East Asia and Pacific (85,264), and Latin America and Caribbean (124,046). Full data is available in **Table S11** in the Supporting Information.

Filtration (i.e., using ceramic, sand, or composite filters) is most prevalent in urban Asian regions (16.1 - 19.6%) and far less common in both rural and urban African regions (0.2% - 3.4%). In contrast, household chlorination appears to be very low in the Asian regions (< 1.5 %) but much more common in Latin America and Caribbean (14.1% [13.8, 14.4] rural; 7.7% [7.5, 7.9] urban) and in the African regions (4.1% - 8.9%) (**Fig. 4, Table S11)**. Thus, future efforts to implement passive chlorinators in the largest target markets described in **Fig. 3** (i.e., rural South Asia and rural East Asia and Pacific) should consider user acceptability and familiarity with chlorine use for drinking water treatment purposes. However, in contrast to manual chlorination methods, passive chlorinators have the potential to be more accepted by households and communities because they eliminate the need for users to manually dose chlorine (thereby limiting user exposure to highly concentrated chlorine solutions) and many technologies can consistently and accurately dose chlorine under taste and odor thresholds.^3,4,23^

To realize the full global potential of passive chlorinators, future research must demonstrate successful and consistent device performance with potentially compatible drinking water sources (including tubewells, boreholes, and protected springs) and solutions that address chlorine supply challenges (e.g., decentralized production via electrochlorination). Our target market estimates could also be further refined and validated by collecting data on the accessibility and usage of storage tanks (compatible with passive chlorinators) at various water points. Lastly, it is critical to develop financially sustainable business models and key implementation partners (NGOs, utilities, and local governments) to effectively scale up passive chlorination in communities and public institutions such as schools and healthcare facilities.

## Data Availability

All data used in this study is publicly available at https://mics.unicef.org/surveys and https://washdata.org/data.

## Acknowledgements

We thank Tom Slaymaker and Richard P. Johnston for their assistance in providing data from the Joint Monitoring Program and for providing critical feedback in reviewing this manuscript. Dr. Katya Cherukumilli was supported by a grant from Open Philanthropy during the writing of this manuscript.

## References

(1) Bain, R.; Johnston, R.; Khan, S.; Hancioglu, A.; Slaymaker, T. Monitoring Drinking Water Quality in Nationally Representative Household Surveys in Low- and Middle-Income Countries: Cross-Sectional Analysis of 27 Multiple Indicator Cluster Surveys 2014-2020. Environ. Health Perspect. 2021, 129 (9), 97010. https://doi.org/10.1289/EHP8459.

(2) Geneva: World Health Organization (WHO) and the United Nations Children’s Fund (UNICEF). Progress on Household Drinking Water, Sanitation and Hygiene 2000-2020: Five Years into the SDGs; WHO & UNICEF, 2021.

(3) Pickering, A. J.; Crider, Y.; Amin, N.; Bauza, V.; Unicomb, L.; Davis, J.; Luby, S. P. Differences in Field Effectiveness and Adoption between a Novel Automated Chlorination System and Household Manual Chlorination of Drinking Water in Dhaka, Bangladesh: A Randomized Controlled Trial. PLoS One 2015, 10 (3), e0118397. https://doi.org/10.1371/journal.pone.0118397.

(4) Lindmark, M.; Cherukumilli, K.; Crider, Y. S.; Marcenac, P.; Lozier, M.; Voth-Gaeddert, L.; Lantagne, D. S.; Mihelcic, J. R.; Zhang, Q. M.; Just, C.; Pickering, A. J. Passive In-Line Chlorination for Drinking Water Disinfection: A Critical Review. Environ. Sci. Technol. 2022. https://doi.org/10.1021/acs.est.1c08580.

(5) Dössegger, L.; Tournefier, A.; Germann, L.; Gärtner, N.; Huonder, T.; Etenu, C.; Wanyama, K.; Ouma, H.; Meierhofer, R. Assessment of Low-Cost, Non-Electrically Powered Chlorination Devices for Gravity-Driven Membrane Water Kiosks in Eastern Uganda. Waterlines 2021, 40 (2), 92–106. https://doi.org/10.3362/1756-3488.20-00014.

(6) Taflin, C. A Low-Cost Solution to Rural Water Disinfection. IEEE Eng. Med. Biol. Mag. 2006, 25 (3), 36–37. https://doi.org/10.1109/MEMB.2006.1636349.

(7) Orner, K. D.; Calvo, A.; Zhang, J.; Mihelcic, J. R. Effectiveness of in-Line Chlorination in a Developing World Gravity-Flow Water Supply. Waterlines 2017, 36 (2), 167–182.

(8) Powers, J. E.; McMurry, C.; Gannon, S.; Drolet, A.; Oremo, J.; Klein, L.; Crider, Y.; Davis, J.; Pickering, A. J. Design, Performance, and Demand for a Novel in-Line Chlorine Doser to Increase Safe Water Access. npj Clean Water 2021, 4 (1), 1–8. https://doi.org/10.1038/s41545-020-00091-1.

(9) Blair, S.; Jooste, D.; Bryant, D.; Ashkar, C.; Burt, S.; Ngo, T. T.; Wolf, D.; Kuwahara, K.; Peterson, J. Humanitarian Engineering Opportunities and Challenges in Rural Dominican Republic: A Case Study of El Cercado. In 2016 IEEE Global Humanitarian Technology Conference (GHTC); 2016; pp 709–716. https://doi.org/10.1109/GHTC.2016.7857356.

(10) Ali, S. I.; Ali, S. S.; Fesselet, J.-F. Effectiveness of Emergency Water Treatment Practices in Refugee Camps in South Sudan. Bull. World Health Organ. 2015, 93 (8), 550–558. https://doi.org/10.2471/BLT.14.147645.

(11) Smith, D. W.; Sultana, S.; Crider, Y. S.; Islam, S. A.; Swarthout, J. M.; Goddard, F. G. B.; Rabbani, A.; Luby, S. P.; Pickering, A. J.; Davis, J. Effective Demand for In-Line Chlorination Bundled with Rental Housing in Dhaka, Bangladesh. Environ. Sci. Technol. 2021, 55 (18), 12471–12482. https://doi.org/10.1021/acs.est.1c01308.

(12) Rayner, J.; Yates, T.; Joseph, M.; Lantagne, D. Sustained Effectiveness of Automatic Chlorinators Installed in Community-Scale Water Distribution Systems during an Emergency Recovery Project in Haiti. Journal of Water, Sanitation and Hygiene for Development. 2016, pp 602–612. https://doi.org/10.2166/washdev.2016.068.

(13) Henderson, A. K.; Sack, R. B.; Toledo, E. A Comparison of Two Systems for Chlorinating Water in Rural Honduras. J. Health Popul. Nutr. 2005, 23 (3), 275–281.

(14) Ngo, T. T.; Medina, J.; White, D.; Jooste, D.; Jerman, K.; Hagen, J.; Peterson, J. Design and Implementation of an Alternative, Low-Cost Water Chlorination System in El Cercado, Dominican Republic. International Journal of Modern Engineering 2018, 13.

(15) Sikder, M.; String, G.; Kamal, Y.; Farrington, M.; Rahman, A. S.; Lantagne, D. Effectiveness of Water Chlorination Programs along the Emergency-Transition-Post-Emergency Continuum: Evaluations of Bucket, in-Line, and Piped Water Chlorination Programs in Cox’s Bazar. Water Res. 2020, 178, 115854.

(16) Amin, N.; Crider, Y. S.; Unicomb, L.; Das, K. K.; Gope, P. S.; Mahmud, Z. H.; Islam, M. S.; Davis, J.; Luby, S. P.; Pickering, A. J. Field Trial of an Automated Batch Chlorinator System at Shared Water Points in an Urban Community of Dhaka, Bangladesh. J. Water Sanit. Hyg. Dev. 2016, 6 (1), 32–41. https://doi.org/10.2166/washdev.2016.027.

(17) Kumpel, E.; Nelson, K. L. Intermittent Water Supply: Prevalence, Practice, and Microbial Water Quality. Environ. Sci. Technol. 2016, 50 (2), 542–553. https://doi.org/10.1021/acs.est.5b03973.

(18) Tools - UNICEF MICS. https://mics.unicef.org/tools (accessed 2021-09-22).

(19) Rosa, G.; Clasen, T. Estimating the Scope of Household Water Treatment in Low- and Medium-Income Countries. The American Journal of Tropical Medicine and Hygiene. 2010, pp 289–300. https://doi.org/10.4269/ajtmh.2010.09-0382.

(20) Methods. https://washdata.org/monitoring/methods (accessed 2021-09-19).

(21) World Population Prospects - Population Division - United Nations. https://population.un.org/wpp/ (accessed 2021-09-19).

(22) Amrose, S. E.; Cherukumilli, K.; Wright, N. C. Chemical Contamination of Drinking Water in Resource-Constrained Settings: Global Prevalence and Piloted Mitigation Strategies. Annu. Rev. Environ. Resour. 2020, 45 (1), 195–226. https://doi.org/10.1146/annurev-environ-012220-105152.

(23) Crider, Y.; Sultana, S.; Unicomb, L.; Davis, J.; Luby, S. P.; Pickering, A. J. Can You Taste It? Taste Detection and Acceptability Thresholds for Chlorine Residual in Drinking Water in Dhaka, Bangladesh. Sci. Total Environ. 2018, 613-614, 840–846. https://doi.org/10.1016/j.scitotenv.2017.09.135.

