## Supplemental Information for "Estimating the Global Target Market for Passive Chlorination"


| Table of Contents Description | Pages |
|---|---|
| Table S1. Joint Monitoring Program criteria for defining safely managed drinking water sources | 2 |
| Table S2: Regional grouping of countries included in market analysis | 3 - 4 |
| Table S3: Compatibility of household drinking water sources with passive chlorinators | 4 |
| Figure S1: Percent (%) of regional populations using household drinking water sources, by facility type (MICS 5 & 6 Surveys) | 5 |
| Table S4: Percent (%) of regional populations using household drinking water sources, by facility type (MICS 5 & 6 Surveys) | 6 |
| Table S5: Percent (%) of regional populations using ***contaminated*** household drinking water sources, by facility type (MICS 5 & 6 Surveys) | 7 |
| Table S6: Percent (%) of regional populations using household drinking water sources that are compatible, potentially compatible, or incompatible with passive chlorinators (JMP 2021 WASH Data) | 8 |
| Table S7: Percent (%) of regional populations using ***contaminated*** household drinking water sources that are compatible, potentially compatible, or incompatible with passive chlorinators (MICS 5 & 6 Surveys) | 9 |
| Table S8: Population (millions) using household drinking water sources, by regional location and source compatibility with passive chlorinators (JMP 2021 WASH Data) | 10 |
| Table S9: Population (millions) using ***contaminated*** household drinking water sources, by regional location and source compatibility with passive chlorinators (JMP 2021 WASH Data and MICS 5 & 6 Surveys) | 11 |
| Table S10: Study limitations and sources of bias | 12 |
| Table S11: Percent (%) of total reporting households using drinking water treatment methods (MICS 5 & 6 Surveys) | 13 |

**Table S1.** Joint Monitoring Program Criteria Defining Safely Managed Drinking Water Sources

| **Source Criteria**[a] | **Criteria Definition** |
|---|---|
| Improved[b] | improved sources include: piped household connections, public taps on standpipes, packaged or delivered water (i.e., tanker trucks and bottled water), rainwater, boreholes, tube wells, protected dug wells, and protected springs |
| Accessible | drinking water sources are located on premises (supply within dwelling, plot, or yard) |
| Available | drinking water sources provide sufficient quantities of water when needed for at least 12 hours/day, 4 days/week |
| Clean | drinking water is free of microbiological contamination (0 *E. coli* CFU/100mL) and priority chemical contamination (< 10 ppb arsenic, and < 1.5 ppm fluoride) |

[a] All criteria must be met for drinking water sources to be classified as “safely managed”
[b] Unimproved sources include unprotected dug wells, unprotected springs, and surface water

**Table S2.** Regional grouping of countries included in analysis

| **LMICs by UNICEF Programme Regions**[a] ($n_{countries}$ = 135) | **Countries with Water Quality Data in MICS 5&6 Databases**[b] ($n_{countries}$ = 37) | **Countries with Water Treatment Data in MICS 5&6 Databases**[b] ($n_{countries}$ = 54 ) |
|---|---|---|
| West and Central Africa (n = 24) | Central African Republic<br>Chad<br>Congo<br>Cote d'Ivoire<br>DRCongo<br>Gambia<br>Ghana<br>Guinea-Bissau<br>Nigeria<br>Sao Tome and Principe<br>Sierra Leone<br>Togo | Benin<br>Cameroon<br>Central African Republic<br>Chad<br>Congo<br>Cote d'Ivoire<br>DRCongo<br>Gambia<br>Ghana<br>Guinea<br>Guinea-Bissau<br>Mali<br>Mauritania<br>Nigeria<br>Sao Tome and Principe<br>Sierra Leone<br>Togo |
| Eastern and Southern Africa (n = 21) | Lesotho<br>Madagascar<br>Malawi<br>Zimbabwe | Eswatini<br>Lesotho<br>Madagascar<br>Malawi<br>Zimbabwe |
| Middle East and North Africa (n = 14) | Algeria<br>Iraq<br>Palestine<br>Tunisia | Algeria<br>Iraq<br>Palestine<br>Sudan<br>Tunisia |
| South Asia (n = 8) | Bangladesh<br>Nepal | Bangladesh<br>Nepal |
| Europe and Central Asia (n = 20) | Georgia<br>Kosovo | Belarus<br>Georgia<br>Kazakhstan<br>Kosovo<br>Kyrgyzstan<br>Montenegro<br>Serbia<br>Turkmenistan |
| East Asia and the Pacific (n = 22) | Fiji<br>Kiribati<br>LaoPDR<br>Mongolia<br>Samoa | Fiji<br>Kiribati<br>LaoPDR<br>Samoa<br>Thailand |

| | | |
|---|---|---|
| | Tonga<br>Tuvalu<br>Vietnam | Tonga<br>Tuvalu<br>Vietnam |
| Latin America and Caribbean<br>(n = 26) | Dominican Republic<br>Guyana<br>Honduras<br>Paraguay<br>Suriname | Belize<br>Cuba<br>Dominican Republic<br>El Salvador<br>Guyana<br>Honduras<br>Mexico<br>Panama<br>Paraguay<br>Suriname |

[a]Russia is classified as being in the region Europe and Central Asia; Venezuela was previously classified by the World Bank as an upper-middle income country but was not classified in 2022 and is considered an LMIC for this study. Argentina and Turks and Caicos island were classified as high-income countries and not included in this study. [b]Survey data for specific population sub-groups that are not nationally representative were excluded (e.g., Kosovo (roma, ashkali), Pakistan-Punjab). Additionally, certain MICS5 surveyed countries *with* water quality modules were included (Congo, Cote d'Ivoire, Nigeria, Paraguay) and MICS6 surveyed countries *without* water quality modules were excluded (Belarus, Costa Rica, Cuba, Kyrgyzstan, Montenegro, North Macedonia, Serbia, Thailand, Turkmenistan).

**Table S3.** Compatibility of Drinking Water Sources with Passive Chlorinators

| **Passive Chlorinator Compatibility** | **Drinking Water Source Types** |
|---|---|
| Compatible Sources | piped water (to dwelling, yard/plot, neighbor, public), kiosks |
| Potentially Compatible Sources | tubewells/boreholes, rainwater, tanker-trucks, cart with small tanks, bottled water and sachet water, protected springs |
| Incompatible Sources | protected and unprotected dug wells, unprotected springs, surface water, other or unclassified sources |

We extracted regional drinking water source data from the 2020 JMP WASH database to classify sources as being "compatible" (piped supplies + water kiosks); "potentially compatible" (improved non-piped sources - protected dugwells); and "incompatible" (unimproved sources + protected dugwells) with passive chlorinators. Using the facility type data in the 2020 JMP WASH database, we estimated the populations using water kiosks (classified as compatible sources) and protected dug wells (classified as incompatible sources) based on countries where these could be clearly identified from the original definitions in the underlying data. Regional mean proportions were used to impute the urban and rural populations using protected dug wells in 93 countries. Owing to the limited number of countries with data on water kiosks, this adjustment was only made for the 16 countries with available data.

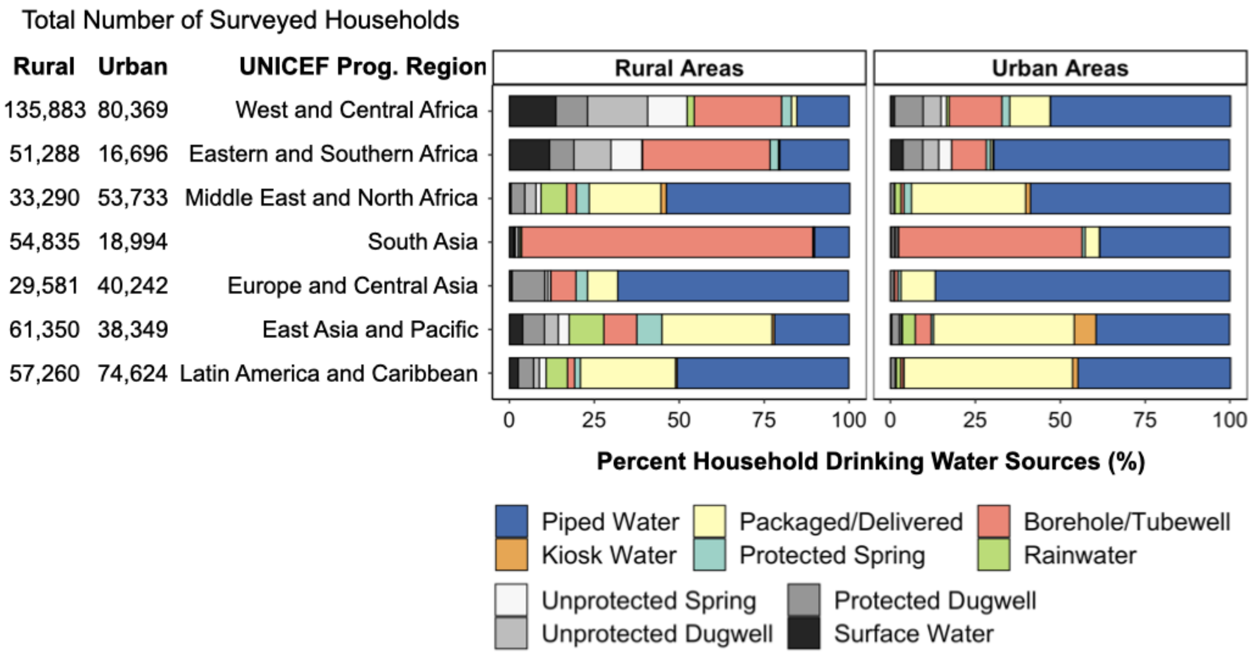


**Figure S1**. Percent (%) of regional populations using household drinking water sources that are compatible, potentially compatible, or incompatible with passive chlorinators, by facility type (MICS 5 & 6 Surveys). This analysis was conducted using the same country surveys with water treatment data (Table S2 column 2), and four additional MICS6 countries with source data (i.e., Costa Rica, Mongolia, North Macedonia, and Serbia). The ratios shown in Fig. S1 are different than the ratios in Fig. 2A, which were calculated using JMP WASH data collected from 135 countries.

**Table S4: Percent (%) of regional populations using household drinking water sources, by facility type (MICS5 & MICS6 Surveys)**

| Water Source Compatability with Passive Chlorinators | West and Central Africa | Eastern and Southern Africa | Middle East and North Africa | South Asia | Europe and Central Asia | East Asia and Pacific | Latin America and Caribbean |
|---|---|---|---|---|---|---|---|
| Rural Areas | | | | | | | |
| Piped Water | 15.4 [ 15.2 , 15.6 ] | 20.3 [ 19.9 , 20.6 ] | 54 [ 53.4 , 54.5 ] | 10.2 [ 10 , 10.5 ] | 67.9 [ 67.4 , 68.5 ] | 22 [ 21.7 , 22.4 ] | 50.5 [ 50.1 , 51 ] |
| Kiosk Water | 0 [ 0 , 0 ] | 0 [ 0 , 0.1 ] | 1.6 [ 1.5 , 1.8 ] | 0.1 [ 0.1 , 0.1 ] | NA | 0.7 [ 0.6 , 0.8 ] | 0.5 [ 0.4 , 0.5 ] |
| Packaged/Delivered | 1.6 [ 1.6 , 1.7 ] | 0.4 [ 0.4 , 0.5 ] | 21.1 [ 20.6 , 21.5 ] | 0.1 [ 0.1 , 0.1 ] | 8.9 [ 8.6 , 9.3 ] | 32.4 [ 32 , 32.7 ] | 28 [ 27.7 , 28.4 ] |
| Protected Spring | 2.9 [ 2.8 , 3 ] | 2.5 [ 2.3 , 2.6 ] | 3.8 [ 3.6 , 4 ] | 0.3 [ 0.2 , 0.3 ] | 3.4 [ 3.2 , 3.6 ] | 7.4 [ 7.2 , 7.6 ] | 1.7 [ 1.6 , 1.8 ] |
| Borehole/Tubewell | 25.7 [ 25.4 , 25.9 ] | 37.5 [ 37.1 , 38 ] | 2.8 [ 2.6 , 3 ] | 85.7 [ 85.4 , 86 ] | 7.4 [ 7.1 , 7.7 ] | 9.7 [ 9.4 , 9.9 ] | 2.1 [ 2 , 2.3 ] |
| Rainwater | 2.1 [ 2.1 , 2.2 ] | 0.2 [ 0.2 , 0.2 ] | 7.6 [ 7.3 , 7.9 ] | 0.5 [ 0.5 , 0.6 ] | 0.1 [ 0.1 , 0.2 ] | 10.2 [ 9.9 , 10.4 ] | 6.3 [ 6.1 , 6.5 ] |
| Unprotected Spring | 11.6 [ 11.4 , 11.7 ] | 9.1 [ 8.9 , 9.4 ] | 1.5 [ 1.3 , 1.6 ] | 0.5 [ 0.4 , 0.5 ] | 0.8 [ 0.7 , 0.9 ] | 3.2 [ 3.1 , 3.4 ] | 2 [ 1.9 , 2.1 ] |
| Unprotected Dugwell | 17.7 [ 17.5 , 17.9 ] | 10.9 [ 10.7 , 11.2 ] | 3.3 [ 3.1 , 3.5 ] | 0.9 [ 0.8 , 1 ] | 0.9 [ 0.8 , 1.1 ] | 4.1 [ 4 , 4.3 ] | 1.7 [ 1.6 , 1.8 ] |
| Protected Dugwell | 9.3 [ 9.2 , 9.5 ] | 7.2 [ 7 , 7.4 ] | 4 [ 3.8 , 4.2 ] | 0.4 [ 0.3 , 0.4 ] | 9.6 [ 9.3 , 9.9 ] | 6.4 [ 6.2 , 6.6 ] | 4.5 [ 4.3 , 4.6 ] |
| Surface Water | 13.7 [ 13.5 , 13.9 ] | 11.8 [ 11.6 , 12.1 ] | 0.5 [ 0.4 , 0.6 ] | 1.3 [ 1.3 , 1.4 ] | 0.8 [ 0.7 , 0.9 ] | 3.9 [ 3.7 , 4.1 ] | 2.6 [ 2.4 , 2.7 ] |
| Urban Areas | | | | | | | |
| Piped Water | 53.1 [ 52.7 , 53.4 ] | 69.5 [ 68.8 , 70.2 ] | 58.9 [ 58.4 , 59.3 ] | 38.4 [ 37.7 , 39.1 ] | 86.8 [ 86.5 , 87.1 ] | 39.4 [ 39 , 39.9 ] | 45 [ 44.7 , 45.4 ] |
| Kiosk Water | 0.1 [ 0.1 , 0.1 ] | 0.3 [ 0.2 , 0.4 ] | 1.4 [ 1.3 , 1.5 ] | 0.1 [ 0.1 , 0.2 ] | NA | 6.4 [ 6.2 , 6.7 ] | 1.6 [ 1.5 , 1.7 ] |
| Packaged/Delivered | 11.9 [ 11.7 , 12.1 ] | 0.7 [ 0.6 , 0.8 ] | 33.6 [ 33.2 , 34 ] | 4.2 [ 3.9 , 4.5 ] | 10.1 [ 9.8 , 10.4 ] | 41.4 [ 40.9 , 41.9 ] | 49.6 [ 49.2 , 49.9 ] |
| Protected Spring | 2.3 [ 2.2 , 2.4 ] | 1.2 [ 1 , 1.3 ] | 2.2 [ 2.1 , 2.4 ] | 0.9 [ 0.7 , 1 ] | 0.9 [ 0.8 , 1 ] | 0.7 [ 0.6 , 0.8 ] | 0.2 [ 0.1 , 0.2 ] |
| Borehole/Tubewell | 15.5 [ 15.3 , 15.8 ] | 10.2 [ 9.8 , 10.7 ] | 0.9 [ 0.8 , 1 ] | 54 [ 53.3 , 54.8 ] | 1 [ 0.9 , 1.1 ] | 4.7 [ 4.5 , 4.9 ] | 0.8 [ 0.7 , 0.9 ] |
| Rainwater | 0.8 [ 0.7 , 0.9 ] | 0 [ 0 , 0.1 ] | 1.8 [ 1.7 , 1.9 ] | 0.2 [ 0.1 , 0.3 ] | 0 [ 0 , 0 ] | 3.8 [ 3.6 , 4 ] | 1.3 [ 1.2 , 1.3 ] |
| Unprotected Spring | 1.6 [ 1.5 , 1.7 ] | 3.8 [ 3.5 , 4.1 ] | 0.1 [ 0.1 , 0.1 ] | 0.6 [ 0.5 , 0.7 ] | 0.1 [ 0.1 , 0.1 ] | 0.3 [ 0.2 , 0.3 ] | 0.1 [ 0.1 , 0.1 ] |
| Unprotected Dugwell | 5.3 [ 5.1 , 5.4 ] | 4.7 [ 4.4 , 5.1 ] | 0.1 [ 0.1 , 0.1 ] | 0.5 [ 0.4 , 0.6 ] | 0.1 [ 0.1 , 0.1 ] | 0.6 [ 0.5 , 0.7 ] | 0.2 [ 0.1 , 0.2 ] |
| Protected Dugwell | 8.4 [ 8.2 , 8.5 ] | 5.8 [ 5.4 , 6.2 ] | 1.1 [ 1 , 1.2 ] | 0.8 [ 0.6 , 0.9 ] | 1 [ 0.9 , 1.1 ] | 2.2 [ 2.1 , 2.4 ] | 1.3 [ 1.2 , 1.4 ] |
| Surface Water | 1.2 [ 1.1 , 1.2 ] | 3.7 [ 3.4 , 4 ] | 0 [ 0 , 0 ] | 0.3 [ 0.2 , 0.4 ] | 0 [ 0 , 0 ] | 0.4 [ 0.3 , 0.5 ] | 0.1 [ 0.1 , 0.1 ] |

95% confidence intervals calculated assuming an underlying binomial distribution are shown in the parentheses.

**Table S5: Percent (%) of regional populations using contaminated household drinking water sources, by facility type (data sources: MICS5 & MICS6 Surveys)**

| Drinking Water Source | West and Central Africa | Eastern and Southern Africa | Middle East and North Africa | South Asia | Europe and Central Asia | East Asia and Pacific | Latin America and Caribbean |
|---|---|---|---|---|---|---|---|
| Rural Areas | | | | | | | |
| Piped Water | 53.7 [ 49.8 , 57.5 ] | 67.6 [ 63.2 , 71.7 ] | 20.5 [ 17.9 , 23.2 ] | 79.5 [ 76.7 , 82.2 ] | 41 [ 38.4 , 43.6 ] | 49.4 [ 46.6 , 52.3 ] | 77.9 [ 75.9 , 79.8 ] |
| Kiosk Water | NA | NA | 82.1 [ 66.5 , 92.5 ] | NA | NA | 6 [ 2 , 13.5 ] | 34.9 [ 25.9 , 44.8 ] |
| Packaged/Delivered | 51.4 [ 34.4 , 68.1 ] | 90.9 [ 58.7 , 99.8 ] | 25.5 [ 20.6 , 31 ] | NA | 3.4 [ 0.1 , 17.8 ] | 30.6 [ 26.5 , 35 ] | 38.7 [ 36.3 , 41.2 ] |
| Protected Spring | 61.3 [ 49.7 , 71.9 ] | 74.5 [ 64.9 , 82.6 ] | 33.1 [ 25.8 , 41.1 ] | 100 [ 89.1 , 100 ] | 50 [ 39.1 , 60.9 ] | 84.5 [ 80.7 , 87.8 ] | 87.8 [ 73.8 , 95.9 ] |
| Borehole/Tubewell | 59.4 [ 56.7 , 62 ] | 53.5 [ 50.6 , 56.5 ] | 21 [ 15 , 28.1 ] | 37.1 [ 35.7 , 38.5 ] | 31.2 [ 20.2 , 44.1 ] | 33 [ 29.9 , 36.2 ] | 77.1 [ 62.7 , 88 ] |
| Rainwater | 87 [ 81.5 , 91.3 ] | 82.1 [ 70.8 , 90.4 ] | 82.4 [ 56.6 , 96.2 ] | 64.7 [ 46.5 , 80.3 ] | NA | 79.4 [ 76.8 , 81.9 ] | 81.1 [ 77.1 , 84.7 ] |
| Unprotected Spring | 91.1 [ 87.6 , 93.8 ] | 93.4 [ 91.2 , 95.2 ] | 61.5 [ 31.6 , 86.1 ] | 88.2 [ 76.1 , 95.6 ] | 62.5 [ 40.6 , 81.2 ] | 79.1 [ 71 , 85.7 ] | 87.4 [ 80.1 , 92.8 ] |
| Unprotected Dugwell | 98.7 [ 97.6 , 99.3 ] | 98.6 [ 97.4 , 99.4 ] | 55 [ 31.5 , 76.9 ] | 96.9 [ 89.2 , 99.6 ] | 87.9 [ 76.7 , 95 ] | 80.6 [ 75.1 , 85.3 ] | 88.1 [ 74.4 , 96 ] |
| Protected Dugwell | 90.8 [ 87.9 , 93.3 ] | 93.8 [ 91.1 , 95.9 ] | 45.8 [ 32.7 , 59.2 ] | 75 [ 55.1 , 89.3 ] | 65.9 [ 59.8 , 71.6 ] | 67.5 [ 63.7 , 71.1 ] | 84.4 [ 76.2 , 90.6 ] |
| Surface Water | 97.6 [ 96.1 , 98.6 ] | 97.6 [ 96.4 , 98.5 ] | NA | 91.4 [ 81 , 97.1 ] | NA | 50.6 [ 44.1 , 57.2 ] | 89.2 [ 79.8 , 95.2 ] |
| Urban Areas | | | | | | | |
| Piped Water | 37.7 [ 34.8 , 40.6 ] | 22.6 [ 20 , 25.2 ] | 14.1 [ 12.6 , 15.6 ] | 75.6 [ 72.8 , 78.2 ] | 6.6 [ 5.4 , 8 ] | 16.8 [ 15.2 , 18.6 ] | 49.6 [ 45.5 , 53.7 ] |
| Kiosk Water | 83.3 [ 35.9 , 99.6 ] | NA | 36.2 [ 22.7 , 51.5 ] | NA | NA | 8.1 [ 5.8 , 11 ] | 40.9 [ 33.3 , 48.8 ] |
| Packaged/Delivered | 36.2 [ 29.1 , 43.8 ] | 22.2 [ 2.8 , 60 ] | 16.2 [ 14.2 , 18.4 ] | 63.1 [ 54.6 , 71.1 ] | 4.3 [ 0.9 , 12 ] | 20.4 [ 16.9 , 24.1 ] | 34.5 [ 32.8 , 36.3 ] |
| Protected Spring | 54.5 [ 32.2 , 75.6 ] | 68.4 [ 43.4 , 87.4 ] | 24.4 [ 17 , 33.1 ] | 92.3 [ 74.9 , 99.1 ] | 33.3 [ 11.8 , 61.6 ] | 36.4 [ 20.4 , 54.9 ] | 87.5 [ 47.3 , 99.7 ] |
| Borehole/Tubewell | 66.2 [ 60.3 , 71.7 ] | 41.9 [ 33.8 , 50.3 ] | 11.8 [ 4.4 , 23.9 ] | 43.5 [ 40.7 , 46.3 ] | 37.5 [ 8.5 , 75.5 ] | 14.8 [ 11 , 19.4 ] | 53.8 [ 25.1 , 80.8 ] |
| Rainwater | 94.4 [ 84.6 , 98.8 ] | NA | 75 [ 42.8 , 94.5 ] | NA | NA | 82 [ 76.7 , 86.5 ] | 84.4 [ 75.3 , 91.2 ] |
| Unprotected Spring | 100 [ 79.4 , 100 ] | 95.8 [ 89.6 , 98.8 ] | 57.1 [ 18.4 , 90.1 ] | 91.7 [ 73 , 99 ] | NA | 100 [ 54.1 , 100 ] | NA |
| Unprotected Dugwell | 93.5 [ 87.1 , 97.4 ] | 100 [ 97.6 , 100 ] | NA | 100 [ 79.4 , 100 ] | 64.3 [ 35.1 , 87.2 ] | 77.8 [ 52.4 , 93.6 ] | 66.7 [ 22.3 , 95.7 ] |
| Protected Dugwell | 95.4 [ 91.5 , 97.9 ] | 95.2 [ 90.8 , 97.9 ] | 20 [ 6.8 , 40.7 ] | 86.7 [ 59.5 , 98.3 ] | 70.4 [ 56.4 , 82 ] | 40 [ 31.5 , 49 ] | 74.4 [ 58.8 , 86.5 ] |
| Surface Water | 87.5 [ 61.7 , 98.4 ] | 100 [ 96.8 , 100 ] | NA | NA | NA | 62.5 [ 24.5 , 91.5 ] | 66.7 [ 22.3 , 95.7 ] |

95% confidence intervals calculated assuming an underlying binomial distribution are shown in the parentheses.

**Table S6: Percent (%) of regional populations using household drinking water sources that are compatible, potentially compatible, or incompatible with passive chlorination (data source: JMP 2021 WASH Data)**

| Water Source Compatability with Passive Chlorinators | Global LMICs | West and Central Africa | Eastern and Southern Africa | Middle East and North Africa | South Asia | Europe and Central Asia | East Asia and Pacific | Latin America and Caribbean |
|---|---|---|---|---|---|---|---|---|
| Total Region | | | | | | | | |
| Compatible | 59.1 | 27.2 | 42.2 | 83.7 | 38.7 | 83.2 | 70.2 | 92.2 |
| Potentially Compatible | 27.9 | 40.1 | 27.6 | 12.7 | 53.8 | 12.5 | 15.5 | 3.5 |
| Incompatible | 13.0 | 32.6 | 30.2 | 3.6 | 7.5 | 4.2 | 14.4 | 4.3 |
| Rural Areas | | | | | | | | |
| Compatible | 38.4 | 14.3 | 24.3 | 74.3 | 27.3 | 66.3 | 50.5 | 76.6 |
| Potentially Compatible | 41.5 | 37.6 | 35.2 | 17.9 | 63.5 | 23.9 | 27.4 | 7.5 |
| Incompatible | 20.1 | 48.1 | 40.5 | 7.8 | 9.2 | 9.7 | 22.1 | 15.9 |
| Urban Areas | | | | | | | | |
| Compatible | 78.6 | 41.5 | 74.8 | 89.3 | 60.0 | 93.0 | 84.6 | 95.9 |
| Potentially Compatible | 15.1 | 43.0 | 13.7 | 9.6 | 35.6 | 5.9 | 6.8 | 2.5 |
| Incompatible | 6.3 | 15.5 | 11.4 | 1.1 | 4.4 | 1.1 | 8.7 | 1.6 |

95% confidence intervals calculated assuming an underlying binomial distribution are shown in the parentheses.

Table S7: Percent (%) of regional populations using contaminated household drinking water sources that are compatible, potentially compatible, or incompatible with passive chlorination (data sources: MICS5 & MICS6 Surveys)

| Water Source Compatability with Passive Chlorinators | Global LMICs | West and Central Africa | Eastern and Southern Africa | Middle East and North Africa | South Asia | Europe and Central Asia | East Asia and Pacific | Latin America and Caribbean |
|---|---|---|---|---|---|---|---|---|
| Total Region | | | | | | | | |
| Compatible | 39 [ 36.6 , 41.4 ] | 42.4 [ 39.2 , 45.6 ] | 39.3 [ 36.1 , 42.5 ] | 17.3 [ 15.5 , 19.3 ] | 77.4 [ 74.6 , 80 ] | 16.7 [ 15 , 18.4 ] | 24.7 [ 22.8 , 26.6 ] | 52.1 [ 48.7 , 55.4 ] |
| Potentially Compatible | 44.8 [ 42.6 , 47 ] | 60.6 [ 57.3 , 63.9 ] | 54.8 [ 51.2 , 58.4 ] | 22.8 [ 20.1 , 25.7 ] | 39.7 [ 38 , 41.4 ] | 28.7 [ 22.1 , 36.4 ] | 51.1 [ 49 , 53.2 ] | 41.6 [ 39.7 , 43.5 ] |
| Incompatible | 80.4 [ 76.7 , 83.6 ] | 95.4 [ 94 , 96.5 ] | 96.4 [ 95.5 , 97.2 ] | 47.2 [ 35.9 , 59 ] | 90.4 [ 84.3 , 94.5 ] | 69 [ 62.7 , 74.8 ] | 60.9 [ 56.4 , 65.4 ] | 82.8 [ 75.9 , 88.4 ] |
| Rural Areas | | | | | | | | |
| Compatible | 56.5 [ 53.6 , 59.3 ] | 54 [ 50.2 , 57.8 ] | 67.6 [ 63.3 , 71.8 ] | 22.9 [ 20.3 , 25.7 ] | 79.4 [ 76.5 , 82.1 ] | 41 [ 38.4 , 43.6 ] | 46.6 [ 43.9 , 49.4 ] | 75.5 [ 73.6 , 77.5 ] |
| Potentially Compatible | 45 [ 43.2 , 46.9 ] | 62.6 [ 60.2 , 64.9 ] | 57 [ 54.2 , 59.7 ] | 27.8 [ 24.3 , 31.5 ] | 37.7 [ 36.3 , 39.1 ] | 35.9 [ 28.9 , 43.4 ] | 57 [ 55.2 , 58.9 ] | 49.2 [ 47.1 , 51.4 ] |
| Incompatible | 84.3 [ 81.8 , 86.5 ] | 95.6 [ 94.6 , 96.4 ] | 96.2 [ 95.4 , 96.9 ] | 50.5 [ 40.1 , 60.9 ] | 90 [ 85.1 , 93.8 ] | 69.4 [ 64.2 , 74.2 ] | 68.1 [ 65.4 , 70.7 ] | 86.9 [ 82.9 , 90.3 ] |
| Urban Areas | | | | | | | | |
| Compatible | 30.9 [ 28.8 , 33.1 ] | 37.9 [ 35 , 40.9 ] | 22.5 [ 20 , 25.2 ] | 14.5 [ 13.1 , 16.1 ] | 75.6 [ 72.9 , 78.2 ] | 6.6 [ 5.4 , 8 ] | 15.1 [ 13.7 , 16.6 ] | 47.7 [ 44.1 , 51.3 ] |
| Potentially Compatible | 44.2 [ 40.9 , 47.5 ] | 58.7 [ 54.4 , 62.9 ] | 44.7 [ 37.3 , 52.3 ] | 17.2 [ 15.3 , 19.3 ] | 46.3 [ 43.7 , 49 ] | 11.8 [ 6.1 , 20.2 ] | 33.7 [ 30.9 , 36.6 ] | 36.2 [ 34.5 , 37.9 ] |
| Incompatible | 68.5 [ 61.6 , 74.7 ] | 94.7 [ 91.7 , 96.8 ] | 97.7 [ 96.1 , 98.8 ] | 32.4 [ 17.4 , 50.5 ] | 91.7 [ 81.6 , 97.2 ] | 67.1 [ 54.9 , 77.9 ] | 47.5 [ 39.6 , 55.5 ] | 73.2 [ 59.7 , 84.2 ] |

95% confidence intervals calculated assuming an underlying binomial distribution are shown in the parentheses.

**Table S8: Population (millions) using household drinking water sources, by regional location and source compatibility with passive chlorinators (data source: JMP 2021 WASH Database)**

| Water Source Compatability with Passive Chlorinators | Global LMICs | West and Central Africa | Eastern and Southern Africa | Middle East and North Africa | South Asia | Europe and Central Asia | East Asia and Pacific | Latin America and Caribbean |
|---|---|---|---|---|---|---|---|---|
| Total Region | | | | | | | | |
| Compatible | 3866.3 | 149.5 | 228.7 | 417.7 | 718.5 | 233.7 | 1519.8 | 598.5 |
| Potentially Compatible | 1824.4 | 220.2 | 149.7 | 63.4 | 998.1 | 35.2 | 335.3 | 22.5 |
| Incompatible | 851.4 | 179.0 | 163.8 | 17.8 | 139.8 | 11.9 | 310.9 | 28.1 |
| Rural Areas | | | | | | | | |
| Compatible | 1218.5 | 41.3 | 85.0 | 138.0 | 330.0 | 68.4 | 461.4 | 94.4 |
| Potentially Compatible | 1316.9 | 108.2 | 123.3 | 33.3 | 767.7 | 24.7 | 250.5 | 9.2 |
| Incompatible | 638.2 | 138.6 | 141.9 | 14.5 | 111.1 | 10.0 | 202.5 | 19.7 |
| Urban Areas | | | | | | | | |
| Compatible | 2647.8 | 108.2 | 143.7 | 279.7 | 388.5 | 165.3 | 1058.4 | 504.0 |
| Potentially Compatible | 507.5 | 112.0 | 26.4 | 30.1 | 230.4 | 10.5 | 84.8 | 13.3 |
| Incompatible | 213.2 | 40.4 | 22.0 | 3.3 | 28.8 | 1.9 | 108.5 | 8.5 |

95% confidence intervals calculated assuming an underlying binomial distribution are shown in the parentheses.

Table S9: Population (millions) using contaminated household drinking water sources, by regional location and source compatibility with passive chlorinators (data sources: MICS5 & MICS6 Surveys, JMP 2021 WASH database)

| Water Source Compatability with Passive Chlorinators | Global LMICs | West and Central Africa | Eastern and Southern Africa | Middle East and North Africa | South Asia | Europe and Central Asia | East Asia and Pacific | Latin America and Caribbean |
|---|---|---|---|---|---|---|---|---|
| Total Region | | | | | | | | |
| Compatible | 1506.8 [ 1415.3 , 1599.1 ] | 63.3 [ 58.6 , 68.1 ] | 89.9 [ 82.5 , 97.3 ] | 72.3 [ 64.6 , 80.5 ] | 555.8 [ 535.7 , 574.7 ] | 39 [ 35.1 , 43.1 ] | 374.8 [ 346.9 , 403.6 ] | 311.8 [ 291.7 , 331.8 ] |
| Potentially Compatible | 817.1 [ 776.3 , 858.3 ] | 133.5 [ 126.1 , 140.7 ] | 82.1 [ 76.7 , 87.4 ] | 14.5 [ 12.7 , 16.3 ] | 396.2 [ 379.7 , 412.8 ] | 10.1 [ 7.8 , 12.8 ] | 171.4 [ 164.4 , 178.5 ] | 9.3 [ 8.9 , 9.8 ] |
| Incompatible | 684.3 [ 653.2 , 711.5 ] | 170.6 [ 168.2 , 172.6 ] | 157.9 [ 156.4 , 159.2 ] | 8.4 [ 6.4 , 10.5 ] | 126.4 [ 117.9 , 132.2 ] | 8.2 [ 7.5 , 8.9 ] | 189.4 [ 175.4 , 203.3 ] | 23.3 [ 21.4 , 24.9 ] |
| Rural Areas | | | | | | | | |
| Compatible | 688 [ 653.2 , 722.1 ] | 22.3 [ 20.7 , 23.9 ] | 57.5 [ 53.8 , 61 ] | 31.7 [ 28 , 35.5 ] | 262 [ 252.6 , 270.8 ] | 28 [ 26.3 , 29.9 ] | 215.1 [ 202.4 , 227.9 ] | 71.3 [ 69.5 , 73.1 ] |
| Potentially Compatible | 593 [ 568.8 , 617.3 ] | 67.7 [ 65.2 , 70.2 ] | 70.3 [ 66.9 , 73.6 ] | 9.3 [ 8.1 , 10.5 ] | 289.4 [ 279 , 300 ] | 8.9 [ 7.1 , 10.7 ] | 142.9 [ 138.2 , 147.5 ] | 4.5 [ 4.3 , 4.7 ] |
| Incompatible | 538.1 [ 521.9 , 552.3 ] | 132.4 [ 131.2 , 133.6 ] | 136.5 [ 135.3 , 137.5 ] | 7.4 [ 5.8 , 8.9 ] | 100 [ 94.5 , 104.2 ] | 7 [ 6.4 , 7.4 ] | 137.8 [ 132.4 , 143.1 ] | 17.1 [ 16.3 , 17.7 ] |
| Urban Areas | | | | | | | | |
| Compatible | 818.9 [ 762 , 877 ] | 41 [ 37.9 , 44.2 ] | 32.4 [ 28.8 , 36.2 ] | 40.7 [ 36.6 , 45 ] | 293.8 [ 283.2 , 303.9 ] | 10.9 [ 8.9 , 13.2 ] | 159.6 [ 144.5 , 175.7 ] | 240.4 [ 222.3 , 258.7 ] |
| Potentially Compatible | 224.1 [ 207.5 , 241.1 ] | 65.8 [ 60.9 , 70.5 ] | 11.8 [ 9.8 , 13.8 ] | 5.2 [ 4.6 , 5.8 ] | 106.8 [ 100.8 , 112.8 ] | 1.2 [ 0.6 , 2.1 ] | 28.6 [ 26.2 , 31 ] | 4.8 [ 4.6 , 5 ] |
| Incompatible | 146.1 [ 131.3 , 159.2 ] | 38.2 [ 37 , 39.1 ] | 21.5 [ 21.1 , 21.7 ] | 1.1 [ 0.6 , 1.7 ] | 26.4 [ 23.5 , 28 ] | 1.3 [ 1 , 1.5 ] | 51.6 [ 43 , 60.2 ] | 6.2 [ 5.1 , 7.1 ] |

95% confidence intervals calculated assuming an underlying binomial distribution are shown in the parentheses. JMP data was analyzed using Stata 17 and MICS data analysis was primarily conducted in R (version 4.2.1).

| Table S10. Study limitations and sources of bias | |
|---|---|
| **Study Assumption** | **Effect on estimated target market for passive chlorination** |
| Exclusion of high-income countries from calculation of global target market | Underestimate |
| Calculations based on MICS water quality measurements taken at a single timepoint; assuming contamination remains consistent over time | Underestimate |
| Calculations based on contamination at the point of collection following definitions for safely managed drinking water services. Households using a water source that is free from contamination at the point of collection but not the point of use were excluded in this analysis but could benefit from passive chlorination. | Underestimate |
| Countries with MICS water quality data are representative of regional water quality | Unclear, varies by region |
| Chlorine is readily available globally and the taste and odor of chlorine are accepted among communities | Overestimation |
| Proportions of contaminated drinking water source types calculated using country-specific MICS data are representative of the regional trends in water source type usage estimated by the JMP data (i.e., these two data sets can be combined to estimate target markets) | Unclear, varies by region |

| Table S11: Percent (%) of total reporting households using drinking water treatment methods (MICS5 & MICS6 Surveys) | | | | | | | |
|---|---|---|---|---|---|---|---|
| Household Water Treatment Method | West and Central Africa | Eastern and Southern Africa | Middle East and North Africa | South Asia | Europe and Central Asia | East Asia and Pacific | Latin America and Caribbean |
| Rural Areas | | | | | | | |
| Boiling | 0.9 [ 0.9 , 1 ] | 10.8 [ 10.6 , 11.1 ] | 1.2 [ 1.1 , 1.4 ] | 1.6 [ 1.5 , 1.7 ] | 21.4 [ 20.9 , 21.9 ] | 36.4 [ 36 , 36.8 ] | 11.8 [ 11.6 , 12.1 ] |
| Filtration | 0.4 [ 0.3 , 0.4 ] | 0.2 [ 0.1 , 0.2 ] | 1.3 [ 1.2 , 1.4 ] | 3.2 [ 3 , 3.3 ] | 6.6 [ 6.3 , 6.9 ] | 8.7 [ 8.5 , 8.9 ] | 2.4 [ 2.3 , 2.5 ] |
| Chlorination | 4.1 [ 4 , 4.2 ] | 8.2 [ 8 , 8.4 ] | 4.4 [ 4.2 , 4.6 ] | 0.4 [ 0.3 , 0.4 ] | 1.3 [ 1.2 , 1.5 ] | 0.3 [ 0.2 , 0.3 ] | 14.1 [ 13.8 , 14.4 ] |
| Settling | 1.4 [ 1.3 , 1.4 ] | 0.6 [ 0.6 , 0.7 ] | 8.3 [ 8 , 8.6 ] | 0.6 [ 0.5 , 0.6 ] | 9.2 [ 8.9 , 9.6 ] | 2.5 [ 2.4 , 2.6 ] | 1.1 [ 1 , 1.2 ] |
| Cloth Straining | 6.5 [ 6.4 , 6.6 ] | 1.2 [ 1.1 , 1.3 ] | 2.6 [ 2.4 , 2.8 ] | 1.3 [ 1.2 , 1.4 ] | 0.4 [ 0.4 , 0.5 ] | 2.3 [ 2.2 , 2.4 ] | 3.8 [ 3.7 , 4 ] |
| Solar Disinfection | 0.1 [ 0.1 , 0.1 ] | 0 [ 0 , 0.1 ] | 0.1 [ 0.1 , 0.1 ] | 0.1 [ 0 , 0.1 ] | 0 [ 0 , 0 ] | 0.2 [ 0.2 , 0.3 ] | 0.2 [ 0.2 , 0.2 ] |
| Other Method | 1 [ 1 , 1.1 ] | 28 [ 27.6 , 28.4 ] | 4.1 [ 3.9 , 4.4 ] | 0.1 [ 0.1 , 0.1 ] | 0.9 [ 0.8 , 1.1 ] | 0.4 [ 0.4 , 0.5 ] | 0.9 [ 0.8 , 1 ] |
| Urban Areas | | | | | | | |
| Boiling | 1.6 [ 1.5 , 1.7 ] | 16.5 [ 16 , 17.1 ] | 1.2 [ 1.1 , 1.3 ] | 12.8 [ 12.3 , 13.2 ] | 21.2 [ 20.8 , 21.6 ] | 27.8 [ 27.3 , 28.3 ] | 11 [ 10.7 , 11.2 ] |
| Filtration | 1 [ 0.9 , 1.1 ] | 0.7 [ 0.6 , 0.9 ] | 3.4 [ 3.2 , 3.6 ] | 16.1 [ 15.6 , 16.6 ] | 19.6 [ 19.2 , 20 ] | 16.1 [ 15.7 , 16.5 ] | 3.4 [ 3.3 , 3.6 ] |
| Chlorination | 8.9 [ 8.7 , 9.1 ] | 5.9 [ 5.5 , 6.3 ] | 4.2 [ 4 , 4.3 ] | 0.6 [ 0.5 , 0.8 ] | 0.2 [ 0.2 , 0.3 ] | 0.3 [ 0.2 , 0.4 ] | 7.7 [ 7.5 , 7.9 ] |
| Settling | 1.5 [ 1.4 , 1.6 ] | 0.2 [ 0.2 , 0.3 ] | 2 [ 1.9 , 2.1 ] | 0.3 [ 0.3 , 0.4 ] | 7.7 [ 7.5 , 8 ] | 1.7 [ 1.6 , 1.9 ] | 0.6 [ 0.5 , 0.6 ] |
| Cloth Straining | 3.9 [ 3.8 , 4 ] | 0.4 [ 0.3 , 0.5 ] | 1 [ 0.9 , 1.1 ] | 3.4 [ 3.1 , 3.6 ] | 0.5 [ 0.4 , 0.5 ] | 1.1 [ 1 , 1.2 ] | 1.6 [ 1.5 , 1.7 ] |
| Solar Disinfection | 0.1 [ 0.1 , 0.1 ] | 0 [ 0 , 0.1 ] | 0 [ 0 , 0 ] | 0.2 [ 0.1 , 0.2 ] | 0 [ 0 , 0 ] | 0.4 [ 0.3 , 0.5 ] | 0.1 [ 0.1 , 0.1 ] |
| Other Method | 1 [ 1 , 1.1 ] | 14.3 [ 13.7 , 14.8 ] | 5.1 [ 4.9 , 5.2 ] | 0.5 [ 0.4 , 0.6 ] | 1.1 [ 1 , 1.2 ] | 0.6 [ 0.5 , 0.7 ] | 0.8 [ 0.7 , 0.8 ] |

95% confidence intervals calculated assuming an underlying binomial distribution are shown in the parentheses.
